# Integrating breast tumor homologous recombination deficiency status to aid germline *BRCA1* and *BRCA2* variant classification

**DOI:** 10.1101/2025.06.12.25329237

**Authors:** Cristina Fortuno, Jia Zhang, Lambros T Koufariotis, Georgina Hollway, Scott Wood, John V Pearson, Peter T Simpson, Sunil R Lakhani, Amy E McCart Reed, Heather Thorne, G Bruce Mann, Anita R Skandarajah, Lisa Devereux, Qihong Zhao, Dilanka L De Silva, Geoffrey J Lindeman, Paul A James, Ian Campbell, Amanda B Spurdle, Nicola Waddell

## Abstract

Pathogenic germline variants in certain genes are associated with somatic tumor mutation signatures. The use of somatic tumor mutation data has the potential to improve the identification of true pathogenic variants but remains underexplored. We investigated the integration of tumor homologous recombination (HR) deficiency status as a predictor of pathogenicity for germline *BRCA1* and *BRCA2* variants, building on the established link between HR deficiency and germline pathogenic variants in these genes. We analyzed breast tumor whole-genome sequence and matching germline data from 350 patients across four datasets: Familial Breast Cancer (N=77), The Cancer Genome Atlas (TCGA-BRCA, N=96), the MAGIC study (N=136), and Q-IMPROvE (N=41). A total of 13,364 germline variants (including structural variations) in *BRCA1*, *BRCA2*, and other DNA repair genes (*ATM*, *BARD1*, *BRIP1*, *CHEK2*, *PALB2*, *PTEN*, *RAD51C*, *RAD51D, TP53*) underwent variant curation. Patients were categorized based on germline classification as *BRCA1* positive (N=25), *BRCA2* positive (N=21), and *BRCA1/2* negative (N=149), excluding those with *BRCA1/2* variants of uncertain significance (N=21) and pathogenic variants in other DNA repair genes (N=134). Somatic HR status (deficient or proficient) was predicted using three algorithms: HRDetect, CHORD, and HRDSum. HR-deficient and HR-proficient status were significant predictors of germline *BRCA1/2* pathogenic variant status (positive and negative directions). The CHORD algorithm, which estimates *BRCA1* and *BRCA2* subtype specifically, added precision contributing evidence towards pathogenicity for the corresponding gene. Finally, we assessed CHORD HR predictions for variants of uncertain significance in *BRCA1* and *BRCA2*, and reported their tumor HR status for potential use as additional evidence in variant curation. Analysis across multiple tumor whole-genome sequencing datasets has shown that HR status prediction algorithms can separate profiles for *BRCA1* and *BRCA2* pathogenic variants and provide further evidence at increased weight to aid in the classification of germline *BRCA1* and *BRCA2* variants. Tumor sequencing offers a promising strategy for reducing the uncertainty in germline variant interpretation.

## Introduction

Homologous recombination deficiency (HRD) impairs a cell’s ability to effectively repair DNA double-strand breaks and underlies tumorigenesis in a significant proportion of breast, ovarian, prostate, and pancreatic cancers.^1^ Approximately 18-30% of breast cancers have been reported to exhibit HR deficiency.^2–4^ Pathogenic germline variations in the hereditary breast and ovarian cancer susceptibility genes *BRCA1* and *BRCA2*, which are crucial for DNA repair through the HR repair pathway, are strongly associated with HRD at the tumor level.^5^ However, HRD has also been associated to some degree with pathogenic germline variation in other DNA repair genes such as *PALB2*^6,7^ and *RAD51C.*^8^

Somatic mutation signatures linked to HRD in BRCA-associated tumors^9,10,11^ have driven the development of several tools to predict HR status, including HRDSum,^12^ HRDetect,^4^ CHORD,^13^ HRDCNA,^14^ and HRProfiler.^15^ Increasing evidence indicates that a tumor with HRD, regardless of germline pathogenic variant status of the patient, predicts response to poly(adenosine diphosphate-ribose) polymerase (PARP) inhibitor therapy in breast,^16^ ovarian^17^ and prostate cancer^18^ patients. As clinical trial evidence accumulates, tumor genomic profiling is expected to become central to the clinical management of patients at diagnosis. This represents a timely opportunity to evaluate a different clinical use of HRD to guide the management of cancer patients - the value and feasibility of using breast tumor tissue profiles generated at the time of diagnosis to aid *BRCA1/2* germline variant classification.

Despite the growing understanding of inherited genetic variations in cancer genes, and the establishment of ClinGen Variant Curation Expert Panels to develop gene-specific classification guidelines, the number of variants of uncertain significance (VUS) that are not clinically actionable, remains a substantial clinical challenge. The incorporation of many types of evidence in germline variant classification algorithms improves the classification of individual VUS. The use of tumor data to aid variant classification has already been investigated. For example, observing somatic variants that are commonly found in tumors can serve as evidence towards the pathogenicity of the same variant observed in the germline setting^19^. While, tumor histopathology features has been used to predict the pathogenicity of germline variants in several genes, including *BRCA1/2*, *TP53* and mismatch repair genes.^20–23^ In this study, we utilize the known association between HRD and *BRCA1/2*-associated breast cancer to explore the use of tumor HRD status in predicting *BRCA1* and *BRCA2* germline variant pathogenicity.

## Methods

### Cohorts of breast cancer patients

The analysis included whole-genome sequencing (WGS) data of 350 primary breast tumor and blood DNA (germline) samples from patients within four study cohorts (**Supplementary Table 1**): kConFab Familial Breast cohort (referred to as Familial Breast), comprising individuals with a personal or family history suggestive of hereditary breast cancer;^24,7^ TCGA-BRCA, representing a diverse cohort of individuals with various clinical, genomic, and molecular characteristics of breast cancer;^25^ the MAGIC study (MAGIC), including women with invasive or high grade *in situ* breast cancer and unknown germline status;^26^ and Q-IMPROvE, a cohort of breast cancer samples that underwent WGS prior to treatment as part of a pilot study to test the value of using WGS in the neoadjuvant setting.^27^ Case and sample de-identified IDs used were the same for the previously published studies (Familial Breast, TCGA-BRCA and Q-IMPROvE), while the identifiers for the MAGIC cohort underwent a two-step de-identification. All tumor samples consisted of fresh frozen tissue except for the MAGIC cohort, which comprised formalin-fixed paraffin-embedded (FFPE)-derived samples. Samples with less than 20x sequencing coverage in tumor or normal samples were excluded. These datasets were used to call somatic variants in tumors for HR prediction, and germline variants in known breast cancer predisposition genes (*BRCA1*, *BRCA2*, *ATM, BARD1*, *BRIP1*, *CHEK2*, *PALB2*, *PTEN*, *RAD51C*, *RAD51D,* and *TP53*).

### Somatic variant calling

WGS data from tumor and patient-matched germline samples were used to identify somatic single-nucleotide variants (SNVs) and insertions and deletions (indels). Specifically, short-read sequencing reads were aligned to the human genome assembly (GRCh38) using BWA-MEM.^28^ Somatic SNVs were identified using a dual calling strategy using the intersection of the post-filtered output of qSNP^29^ and GATK^30^. Short insertions and deletions (1-50 bp) were detected with GATK. Sequence alterations were annotated with qannotate to identify somatic-specific variants and to filter variants located within 6 base pairs of a homopolymer region, and with SnpEff^31^ for gene consequence.

Somatic copy number aberrations (CNA) were identified using the tool ascatNGS.^32^ The copy number state of each gene was determined by annotation against known Ensembl genes (version 112). Structural variants (SVs) were determined using qSV;^33^ https://github.com/AdamaJava/adamajava) using both tumor and germline alignments and subsequent filtering to include high-confidence calls in the analysis.

### Single-base substitution signature analysis

The Fit function of the R package signature.tools.lib (v2.4.5) was used to estimate the number of single-base-substitution COSMIC (v2) signatures based on the somatic SNV catalogues of each tumor sample with 1000 bootstraps. The proportions of each signature were calculated based on the total number of variants.

### Correction of the FFPE mutational signature in the MAGIC cohort

Genomic analysis of DNA extracted from FFPE-derived samples can be problematic, as formalin fixation negatively impacts DNA quality and quantity compared to fresh frozen material. The MAGIC cohort tumor DNA was derived from FFPE samples, therefore FFPEsig (https://github.com/QingliGuo/FFPEsig) was used to correct the FFPE noise signatures from the observed mutational catalogues within these samples. This was performed using the unrepaired mode (without uracil DNA glycosylase).^34^ The corrected profiles were then used in the signature analysis and for the HR prediction.

### Homologous recombination deficiency predictions

The somatic tumor variant profiles were analyzed for each individual to predict HR status, classified as deficient (HRD) or proficient (HRP) using three prediction methods, CHORD,^13^ HRDetect,^4^ and HRDSum.^12^

The CHORD pan-cancer HR predictor uses a random forest model trained with samples from different cancer types.^13^ CHORD uses the somatic SNVs, indels and SVs as input, and primarily infers HRD from the relative proportions of microhomology-mediated deletions and 1-10kb duplications. Additionally, CHORD utilizes 1-100kb structural duplications to distinguish *BRCA1* subtype HRD from *BRCA2* subtype HRD. A score of >0.5 is considered to represent deficiency.^13^

The HRDetect method uses the mutational signatures to predict *BRCA1/2* deficiency as a surrogate for HR deficiency,^4^ including base substitution signatures 3 and 8, indel patterns, SVs and the copy number-based score HRD loss of heterozygosity (LOH). The HRDetect_pipeline function of signature.tools.lib was used for the HRDetect implementation with 1000 bootstraps and “Breast” specific signatures. A score of >0.7 is considered to represent deficiency.^4^

HRDsum score is a summary score previously used in clinical trials and is based on LOH,^35^ large-scale state transitions^36^ and the number of telomeric allelic imbalances^37^ calculated from the somatic copy number profile.^12^ The HRDsum score was estimated for each sample using the modified scripts of the R package scarHRD.^38^ A cutoff point of 42 is currently considered an US Food and Drug Administration-approved biomarker to select ovarian cancer patients for PARP inhibition,^39^ which we used to differentiate between deficiency and proficiency in this study.

### Germline variant calling and annotation

The germline sequence data were processed with the GATK best practice workflow to detect germline SNVs and small indels. The nanno module of qannotate (https://github.com/AdamaJava/adamajava) was used to annotate SNVs against dbNSFP^40^ (v4.1a), ClinVar^41^, and gnomAD^42^ (v3.1.2). The annotations for the nine breast cancer susceptibility genes (*BRCA1*, *BRCA2*, *ATM, BARD1*, *BRIP1*, *CHEK2*, *PALB2*, *PTEN*, *RAD51C*, *RAD51D,* and *TP53*) were extracted, and unique SNVs and indels were collected across 350 individuals.

SVs in the germline genome were identified using DELLY (0.7.8) for each individual. SVs overlapping with target genes within a 100bp window were extracted for classification. A total of eight SVs in the related genes from six individuals, which were classified as pathogenic based on the Variant Effect Predictor consequence predicted null (i.e. frameshift, stop loss, coding sequence variants involving complex rearrangements in clinically relevant exons), were included for analyses as high-confidence germline variants.

### Germline variant curation and individual groupings

All variants were reported and classified in relation to the MANE transcripts, as follows: NM_007294.4 (*BRCA1*), NM_000059.4 (*BRCA2*), NM_000051.4 (*ATM*), NM_000465.4 (*BARD1*), NM_032043.3 (*BRIP1*), NM_007194.4 (*CHEK2*), NM_024675.4 (*PALB2*), NM_000314.6 (*PTEN*), NM_058216.3 (*RAD51C*), NM_002878.4 (*RAD51D*), and NM_000546.6 (*TP53*). Germline SNVs and indels were classified using a combination of ClinVar lookups and summary data review, filtering allele frequency (FAF) ≥0.0001 in the gnomAD database, variant effect, and bioinformatic prediction of variant impact using BayesDel and maximum SpliceAI delta score. Variants were collapsed into one of three groups – pathogenic/likely pathogenic (P/LP), benign/likely benign (B/LB) or variant of uncertain significance (VUS). Variants in *BRCA1* and *BRCA2* were additionally classified following the ENIGMA *BRCA1* and *BRCA2* Variant Curation Expert Panel (VCEP) specifications v1.1.0.^43^ Each variant was assigned a category related to whether the variant was within *BRCA1/2* or other genes (*ATM, BARD1*, *BRIP1*, *CHEK2*, *PALB2*, *PTEN*, *RAD51C*, *RAD51D,* and *TP53*). The categories were as follows (**Supplementary Table 2**): A: P/LP_Other genes (where suspicious VUS were also conservatively included), B: VUS_Other genes, C: B/LB = Other genes, D: P/LP_BRCA, E: VUS_BRCA, and F: B/LB_BRCA. The eight high-confidence pathogenic germline SVs identified using DELLY (three in *BRCA1*, three in *BRCA2*, one in *PALB2*, and one in *RAD51C*) were included in the relevant germline categories.

Based on these categories and their combinations in the same individual, cases were grouped into four different groups for analysis (**Supplementary Table 3**). These groups included *BRCA1* and *BRCA2* positive groups (individuals carrying a P/LP variant in *BRCA1* or *BRCA2*, respectively) and a *BRCA1/2* negative group (individuals without a detectable *BRCA1/2* P/LP variant). To avoid the confounding effect of pathogenic variants or VUS in the DNA repair genes that may be linked to tumorigenesis, two additional groups were created, termed Excluded (individuals with P/LP or suspicious VUS in other selected DNA repair genes), and BRCA1/2 VUS (individuals with VUS in BRCA1 or BRCA2, and with no other P/LP variants in any genes); both of these additional groups were excluded from the BRCA1/2 positive and negative groups used for main analyses.

### Likelihood ratio calculations

Likelihood ratios (LRs) associated with each HR dichotomous status (HRP vs HRD) were calculated as predicted by each tool (HRDetect, CHORD, and HRDsum), using previously used methods;^20^ this involved comparison of the proportion of HRD-predicted tumors observed for *BRCA1/2* negative individuals compared to that observed for *BRCA1* positive individuals, and separately for *BRCA2* positive individuals. For CHORD-predicted, the LRs were estimated by stratifying HRD status further into *BRCA1* and *BRCA2* HRD subtypes. A sensitivity analysis was performed including individuals in the Excluded groups. The LRs were used to assign an ACMG/AMP evidence strength category and points using Bayesian conversions.^44,45^

### Correlation with histopathology data

We collected tumor histopathology data comprising histological grade and hormone receptor status (ER, PR, HER2, and the combined triple-negative breast cancer, TNBC). Samples with missing histopathology data for a given variable were excluded from corresponding analysis. To examine the association between tumor pathological measures (grade and hormone receptor status), we performed chi-square tests. Effect size was estimated using Cramér’s V to assess the strength of association. Correlations between HR status, as predicted by the different tools, and tumor histopathology markers (grade and hormone receptor status) were analyzed using Cramér’s V to assess the significance of associations. In addition, for CHORD we compared the BRCA1-type HRD probability scores between TNBC and non-TNBC samples using the Wilcoxon rank-sum tests.

## Results

WGS somatic and paired germline data from 350 breast cancer patients within four cohorts were processed **(Supplementary Table 1)**. Three of these cohorts (MAGIC, TCGA-BRCA and Q-IMPROVE) were comprised of unselected breast cancer patients, and the fourth cohort comprised patients with familial cancer. The average tumor purity of samples was 0.58 (Familial breast: 0.56, TCGA-BRCA: 0.60, MAGIC: 0.60 and Q-IMPROvE: 0.46) (**Supplementary Table 1 and Supplementary Figure 1A**). To harmonize the data, all data were re-processed with the same pipeline for somatic variant detection. Samples from the MAGIC cohort yielded fewer somatic variants, with a lower tumor mutation burden (TMB) compared to other cohorts (**Supplementary Figure 1B**) and fewer somatic SV events (p=2.2e-16) (**Supplementary Table 1, Supplementary Figure 1C**), likely due to the impact of FFPE-derived samples on DNA quality and lower read depth in these tumor samples.

### Classification of patients based on *BRCA1/2* and other gene variants

Across the 350 individuals, a total of 13,364 germline SNVs, indels, and SVs were detected in *BRCA1*, *BRCA2* and non-*BRCA1/2* genes, including HR-related genes (*BARD1*, *BRIP1*, *PALB2, PTEN, RAD51C*, *RAD51D,* and *TP53)* as well as *ATM* and *CHEK2*. These germline variants were grouped into six categories as defined in **Figure 1A** and **Supplementary Table 2**. Each individual person was then placed into a germline group based on germline pathogenic variant status for all genes: 25 individuals were classified as *BRCA1* positive, 21 as *BRCA2* positive, 149 as *BRCA1/2* negative, 134 were excluded from analyses (Excluded) as they were identified to have P/LP variants in other non-*BRCA1/2* genes, and 21 were considered for additional analyses (*BRCA1/2* VUS) (**Figure 1B and Figure 1C**).

**Figure 1.**
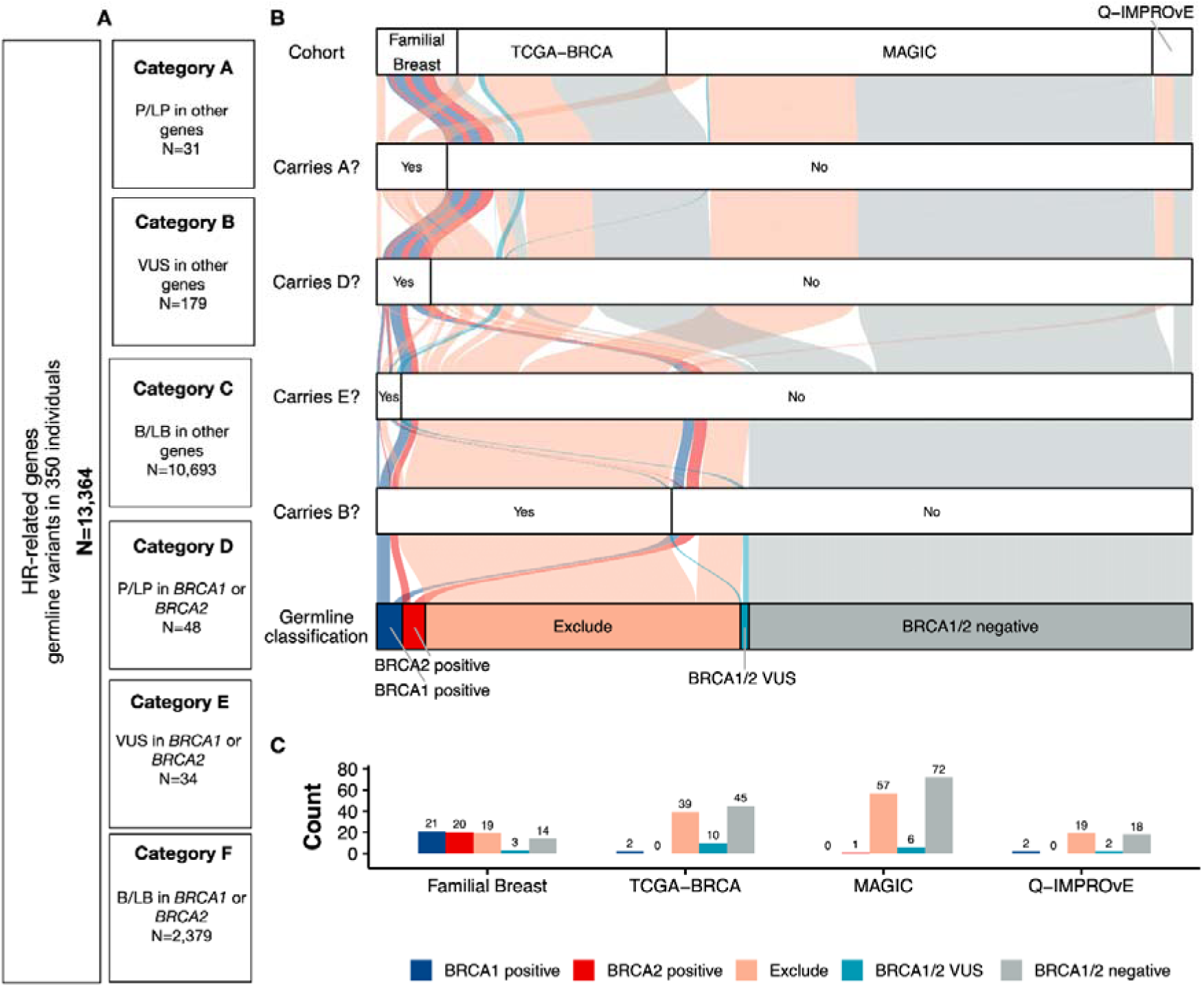
Germline variant classifications and individual allocation into a germline group based on the presence of germline variants. A) The total number of germline variants (including SNVs, indels and SVs) identified in the *BRCA1*, *BRCA2*, *ATM*, *BARD1*, *BRIP1*, *CHEK2, PALB2, PTEN, RAD51C*, *RAD51D* and *TP53* genes. Each variant was classified into six categories (A to F) as defined in the boxes. B) The alluvial diagram illustrates the allocation of 350 individuals into five color-coded germline groups (dark blue: *BRCA1* positive; red: *BRCA2* positive; pink: Exclude; teal: *BRCA1* or *BRCA2* VUS; grey: *BRCA1/2* negative). Individuals were classified into each group based on the four criteria displayed along the y-axis. C) The bar plot displays the individuals classified into five groups within each patient cohort (Familial Breast, TCGA-BRCA, MAGIC and Q-IMPROvE)

### HR profile according to cohort and prediction method

Three methods for HR prediction, based on tumor genomic scars, were used to predict the tumor HR status of the 350 samples. We used the previously published thresholds to determine whether a sample was HRD: CHORD >0.5,^13^ HRDetect >0.7,^4^ and HRDSum >42.^12^ The HRDetect and CHORD predicted HRD probabilities showed distinct separation at the selected thresholds for classifying HR status (**Figure 2A and 2B**), while HRDsum scores had no clear distinction between HRD and HRP breast tumors irrespective of threshold (**Figure 2C**). The two machine-learning-based approaches, HRDetect and CHORD, were highly concordant with each other and consistently identified 104 HRD and 241 HRP samples (**Figure 2D and Supplementary Figure 2**). Only five samples were predicted differently by HRDetect and CHORD. One sample was predicted HRD by CHORD but proficient by HRDetect. This sample was in the Excluded group with a germline LP/P variant in *PALB2* (category A). The other four samples were predicted as HRD by HRDetect but not CHORD: one in the *BRCA1/2* negative group, and three samples in the Excluded group (**Figure 2D, Supplementary Table 4**). The non-congruent HR classification for these samples may be attributed to a small number of SV events in one case (MAGIC37, N=8), or the presence of VUS in other selected DNA repair genes (*PALB2*, *ATM* or *BRIP1*) **Supplementary Table 4**).

**Figure 2.**
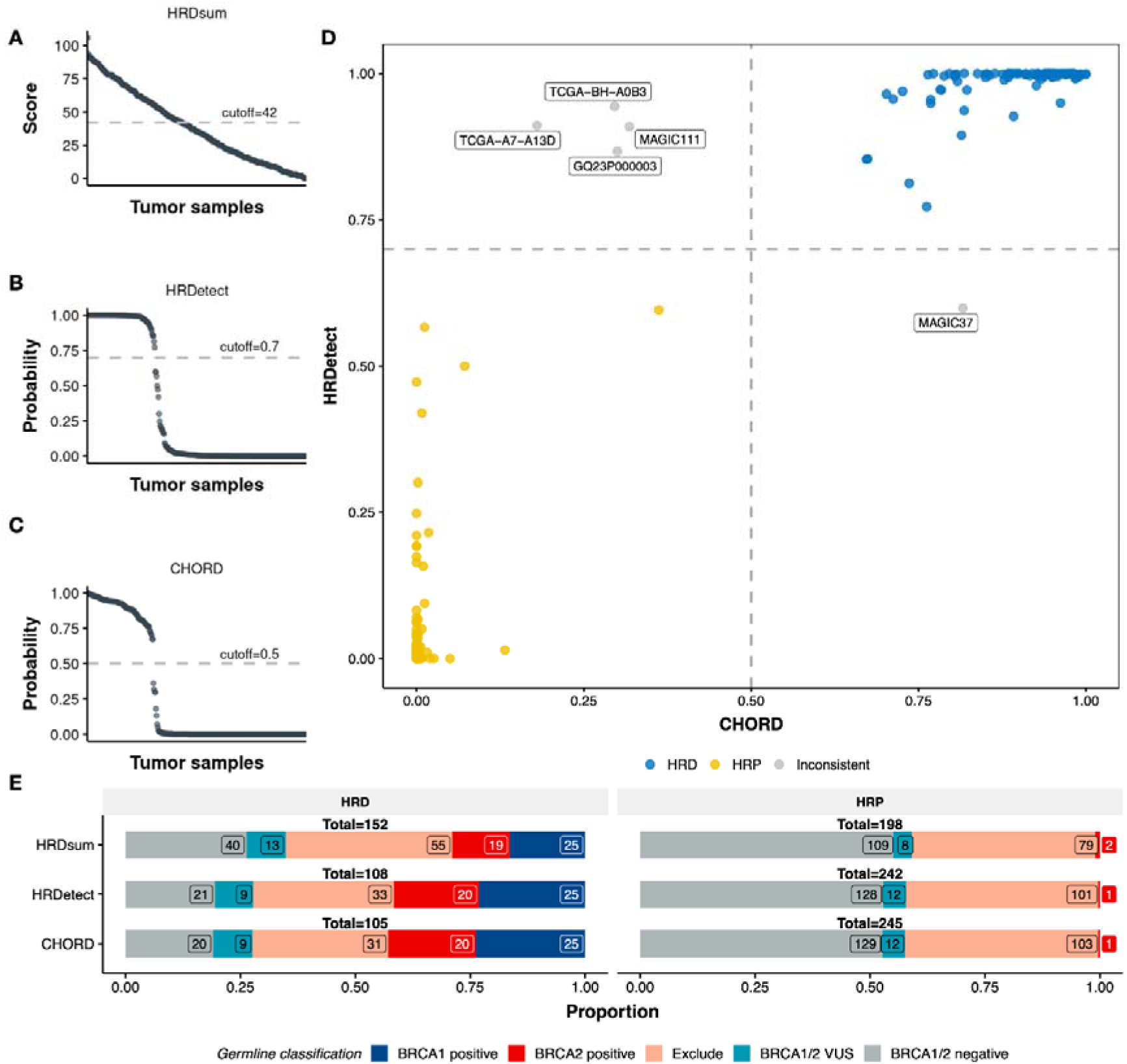
Comparison of HR status predictions using three approaches. The distribution of prediction scores or probabilities for all tumor samples using three methods: **(A)** CHORD, (**B**) HRDetect, and (**C**) HRDsum, with cutoff values indicated as grey dashed lines in the plots. **D**) The comparison of HR prediction between CHORD and HRDetect. HRD samples are labelled as blue points and HRP samples as yellow points. Five samples with inconsistent predictions between CHORD and HRDetect are labelled in grey. **E**) The number of samples predicted as HRD or HRP by the three tools indicated on the y-axis. The bars are colored by the proportion of individuals within each classification group (Dark blue: *BRCA1* positive; red: *BRCA2* positive; pink: Exclude; teal: *BRCA1* or *BRCA2* VUS; grey: *BRCA1/2* negative)

HRDsum identified more HRD samples than HRDetect and CHORD, predicting 40 samples in the *BRCA1/2* negative group as HRD (**Figure 2E**). In comparison, HRDetect and CHORD only predicted 21 and 20 *BRCA1/2* negative samples as HRD, respectively. This suggests that HRDsum may overcall HRD samples, particularly in those non-familial breast cohorts (**Supplementary Figure 3**). All individuals carrying a pathogenic *BRCA1* germline variant were consistently predicted as HRD by all three methods. However, HRDsum predicted two *BRCA2*-positive individuals as HRP, while HRDetect and CHORD only predicted one of them as HRP (**Figure 2E**). Interestingly, this *BRCA2* positive case predicted HRP using all tools did not show evidence of a “second hit” in the tumor, as previously reported,^7^ whereas all other individuals with *BRCA1* or *BRCA2* pathogenic germline variants showed evidence of a somatic event leading to loss of the reference allele, or gain of the variant allele (**Supplementary Table 4**). This suggests that the HR proficient tumor from the *BRCA2* positive individual is likely unrelated to the *BRCA2* germline variant, and due to another unknown mechanism of tumorigenesis.

Overall, the proportion of HRD tumors in each of the four cohorts was consistent with the proportion of *BRCA1/2* positive individuals included in that cohort, being highest in Familial breast where approximately 50% of the individuals carried *BRCA1/2* germline pathogenic variants, and lowest in MAGIC where there was only one *BRCA2* positive individual (**Figure 1C**).

The MAGIC cohort consisted of sequence data from FFPE samples, therefore, we tested whether a signature correction for SNVs would alter the HR status prediction. Although there were minor differences in the score assigned, the prediction of HR status did not change after the signature correction for CHORD (**Supplementary Figure 4A**) or HRDetect (**Supplementary Figure 4B**).

### Prediction of *BRCA1* and *BRCA2*-associated HRD subtype using CHORD

In addition to differentiating between HR deficient and proficient status, the CHORD tool can predict whether an HRD case is likely associated with *BRCA1* or *BRCA2* by providing an HRD subtype. CHORD subtype prediction of the 105 HRD samples identified 60 as *BRCA1* type (32 in Familial breast, 17 in TCGA-BRCA, two in MAGIC, and nine in Q-IMPROvE), and 40 as *BRCA2* type (21 in Familial Breast Cancer, 13 in TCGA-BRCA, five in MAGIC, and one in Q-IMPROvE) (**Figure 3A**). The remaining HRD samples (four in MAGIC and one in Q-IMPROvE), none falling in the *BRCA1/2* positive germline groups, had an undetermined subtype as they could not be assigned to *BRCA1* or *BRCA2* subtype. While some of these undermined samples may be driven by a small number of somatic SV events detected in these samples, the presence of germline or somatic variation in other genes (e.g. *PALB2* in MAGIC32, MAGIC37) may also contribute to the undetermined subtype categorization. Further training of CHORD on HRD cases associated with other genes (not *BRCA1* or *BRCA2*) may improve the prediction of these cases.

**Figure 3.**
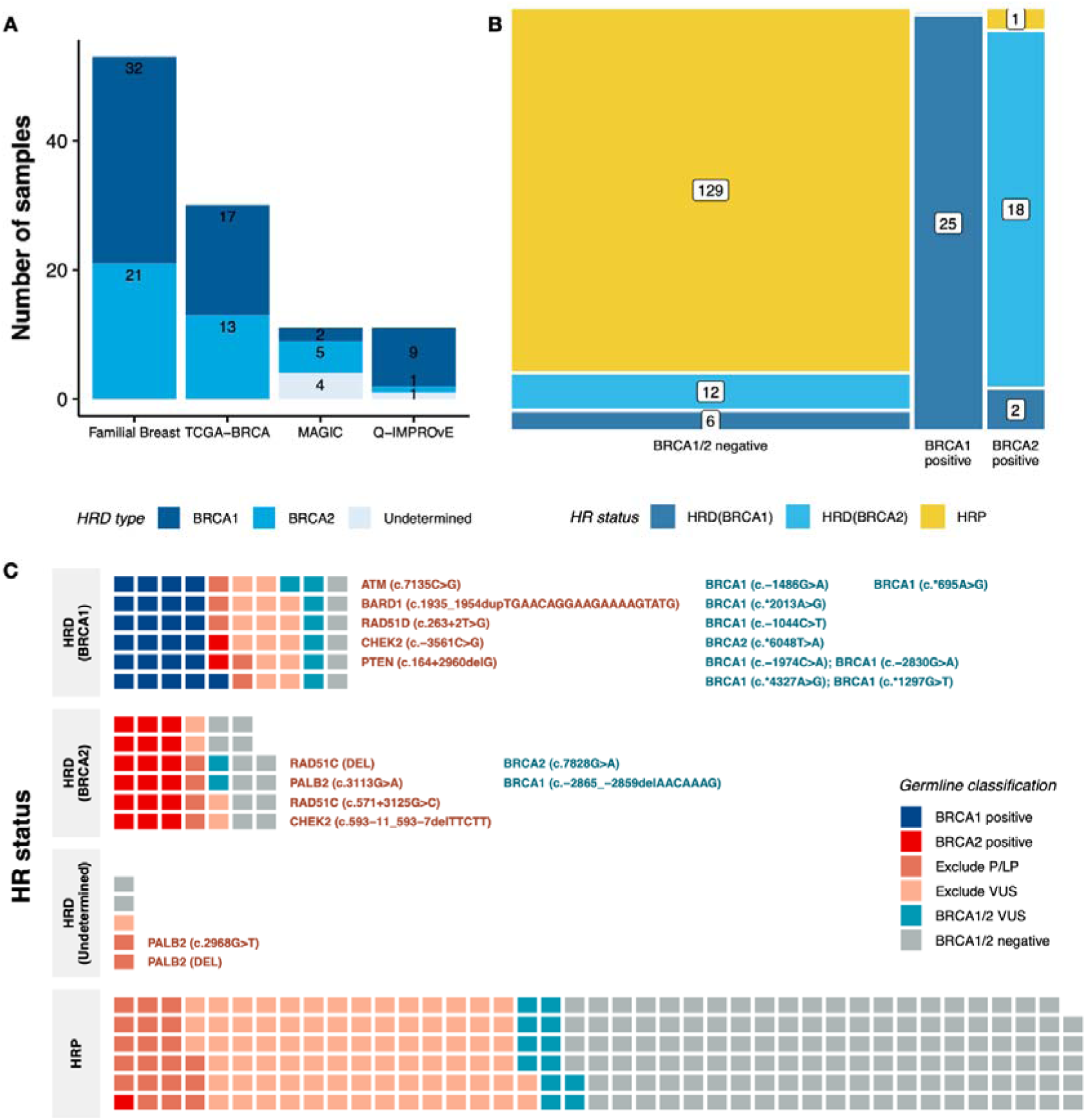
HRD subtype predicted by CHORD and samples from germline classification groups for different HR statuses. **A)** The number of HRD samples differentiated as *BRCA1* and *BRCA2* subtypes or undetermined (five samples). Samples are grouped by the study cohort (Familial breast, TCGA-BRCA, MAGIC and Q-IMPROvE). **B)** A mosaic diagram shows the proportion of HRD samples in the *BRCA1/2* negative group and *BRCA1* or *BRCA2* positive groups. The color indicates the HR status predicted by CHORD (Dark blue HRD *BRCA1* subtype, light blue HRD *BRCA2* subtype, yellow HRP). **C)** Waffle plots indicate the number of individuals assigned to each germline group among the samples characterized as tumor HRD (*BRCA1* subtype, *BRCA2* subtype, undetermined) and HRP. The Excluded group was further divided into those harboring P/LP variants or suspicious VUS in other DNA repair genes. The P/LP variants detected in non-*BRCA1/2* genes and BRCA1/2 VUS of HRD samples are labelled in the plot

All 25 *BRCA1* positive samples were predicted to have a *BRCA1* HRD subtype, and 18 of the 21 *BRCA2* positive samples had a *BRCA2* HRD subtype. For the 149 *BRCA1/2* negative samples, the majority were predicted HRP (n=129, 87%), with the remainder predicted as HRD with an undetermined subtype (n=2), *BRCA1* subtype (n=6), or *BRCA2* subtype (n=12) (**Figure 3B**). A review of the somatic variants for these germline *BRCA1/2* negative samples with *BRCA1* or *BRCA2* HRD subtype predictions revealed relevant somatic changes in these tumors (**Supplementary Table 4**). The six *BRCA1* subtype tumors contained somatic copy number events impacting *BRCA1* (four with copy neutral LOH and two with gains). The 12 *BRCA2* subtype tumors contained somatic copy number events impacting *BRCA2* (two with homozygous loss, four with loss of one allele, four with copy neutral LOH and two with gains). Two of these samples also contained a somatic *BRCA2* splice variant in one case and a somatic inframe *PALB2* variant in another case (**Supplementary Table 4**).

Overall, samples predicted to have HRD *BRCA1* or *BRCA2* subtypes by CHORD, were enriched for individuals that were positive for a germline *BRCA1* or *BRCA2* pathogenic variant, respectively, while *BRCA1/2* negative individuals accounted for more than 50% of the HRP samples (**Figure 3C**). Interestingly, of the 31 Excluded samples with P/LP variants in other DNA repair genes, nine had HRD *BRCA1* or *BRCA2* subtype tumors. These variants are specified in **Figure 3C**.

### HR status as evidence for or against the pathogenicity of *BRCA1* and *BRCA2* germline variants

To determine if CHORD HR status is a significant predictor of *BRCA1* and *BRCA2* germline variant pathogenicity, the proportion of individuals with each HR status was compared for 149 *BRCA1/2* negative individuals to the 25 *BRCA1* positive or the 21 *BRCA2* positive individuals, in order to calculate LRs towards pathogenicity. For *BRCA1*, observation of an HRP breast tumor corresponded to benign strong evidence for that variant (**Table 1**), while observation of an HRD breast tumor corresponded to pathogenic moderate evidence. For *BRCA2*, the strength of HRP status as a predictor against pathogenicity was lower than for *BRCA1*, i.e. benign moderate, while the evidence strength towards pathogenicity associated with CHORD-predicted HR status was the same as derived for *BRCA1* i.e. pathogenic moderate.

**Table 1.** LR calculations associated with HR status predicted by CHORD.

| **BRCA1 - Dichotomous** | **BRCA1/2 negative**<br>**reference set (n)** | **%** | **BRCA1 positive**<br>**reference set (n)** | **%** | **LR (95% CI)** | **Evidence Strength (Points)** |
| --- | --- | --- | --- | --- | --- | --- |
| HRP | 129 | 0.87 | 0 | 0.00 | 0.02 (0.00, 0.35) | Benign Strong (-4) |
| HRD | 20 | 0.13 | 25 | 1.00 | 7.45 (4.96, 11.20) | Pathogenic Moderate (+2) |
| Total | 149 |  | 25 |  |  |  |
| **BRCA1 - Stratified by**<br>**subtype** | **BRCA1/2 negative**<br>**reference set (n)** | **%** | **BRCA1 positive**<br>**reference set (n)** | **%** | **LR (95% CI)** | **Evidence Strength (Points)** |
| HRP | 129 | 0.87 | 0 | 0.00 | 0.02 (0.00, 0.35) | Benign Strong (-4) |
| HRD BRCA1 subtype | 6 | 0.04 | 25 | 1.00 | 24.83 (11.34, 54.38) | Pathogenic Strong (+4) |
| HRD BRCA2 subtype | 12 | 0.08 | 0 | 0.00 | 0.23 (0.01, 3.78) | Not significant (0) |
| Total | 149* |  | 25 |  |  |  |
| **BRCA2 - Dichotomous** | **BRCA1/2 negative**<br>**reference set (n)** | **%** | **BRCA2 positive**<br>**reference set (n)** | **%** | **LR (95% CI)** | **Evidence Strength (Points)** |
| HRP | 129 | 0.87 | 1 | 0.05 | 0.06 (0.01, 0.37) | Benign Moderate (-2) |
| HRD | 20 | 0.13 | 20 | 0.95 | 7.10 (4.67, 10.79) | Pathogenic Moderate (+2) |
| Total | 149 |  | 21 |  |  |  |
| **BRCA2 - Stratified by**<br>**subtype** | **BRCA1/2 negative**<br>**reference set (n)** | **%** | **BRCA2 positive**<br>**reference set (n)** | **%** | **LR (95% CI)** | **Evidence Strength (Points)** |
| HRP | 129 | 0.87 | 1 | 0.05 | 0.06 (0.01, 0.37) | Benign Moderate (-2) |
| HRD BRCA1 subtype | 6 | 0.04 | 2 | 0.10 | 2.37 (0.51, 10.96) | Not significant (0) |
| HRD BRCA2 subtype | 12 | 0.08 | 18 | 0.86 | 10.64 (6.02, 18.82) | Pathogenic Moderate (+2) |
| Total | 149* |  | 21 |  |  |  |

\*Includes two tumors with HRD subtype undetermined

The LR towards pathogenicity increased further for CHORD predictions stratified by the gene-specific HRD subtype (**Table 1**), reaching pathogenic strong evidence for *BRCA1*. Notably, the LR for the HRD subtype inconsistent with the gene being analyzed was not significant (based on the confidence intervals).

Similar results were seen when HRDetect was used to predict HR status regardless of subtype (**Supplementary Table 5**), although the LRs were of slightly lower magnitude in both benign and pathogenic directions using HRDetect (0.02 and 7.10 for *BRCA1*, 0.06 and 6.76 for *BRCA2*) compared to CHORD (0.02 and 7.45 for *BRCA1*, 0.06 and 7.10 for *BRCA2*). HRDSum also resulted in benign strong and moderate evidence for *BRCA1* and *BRCA2*, respectively, but with LRs of the lowest magnitude, while the evidence strength for *BRCA1* and *BRCA2* variants was equivalent only to pathogenic supporting evidence (**Supplementary Table 6**).

We also conducted a sensitivity analysis for the CHORD predictions considering the additional data for 134 individuals previously excluded (as they were identified to have P/LP variants in other non-*BRCA1/2* genes). Among these, three were positive for *BRCA1*, and another two were positive for *BRCA2*. Reanalysis considering prediction of *BRCA1* and *BRCA2* positive status irrespective of presence of pathogenic variants in other genes revealed that LRs did not significantly change and the corresponding evidence categories remained the same, except that HRP status in *BRCA2* increased in strength from benign moderate to benign strong (**Supplementary Table 7**).

### Evaluation of HR predictions in individuals with a BRCA1/2 germline VUS

There was a total of 21 breast cancer patients carrying 23 *BRCA1* or *BRCA2* germline variants classified as VUS that had been excluded from the reference sets used in the LR analyses. All variants were absent from gnomAD except NM_007294.4(BRCA1):c.-60C>T, with two alleles across gnomAD v2 and v3.

Nine of the VUS (n=8 *BRCA1* and n=1 *BRCA2*) identified in seven individuals demonstrated tumor HRD (**Supplementary Table 4**) providing evidence in favor of pathogenicity (**Supplementary Table 8**); however, two individuals each carried two VUS, making it unclear which specific variant could be contributing to the observed HRD. Of the individuals with a *BRCA1* germline VUS with HRD, all had undergone somatic copy number events resulting in bi-allelic loss of *BRCA1* (loss of one allele or copy neutral LOH), providing additional support. Twelve of the VUS (in 12 individuals) had HRP status providing evidence against pathogenicity (**Supplementary Table 8**). The remaining two VUS had HRD subtypes contrasting with the gene in which they were identified (i.e., *BRCA1* VUS with HRD *BRCA2* subtype, and vice versa) – while, statistically, this observation provides no evidence for or against pathogenicity, it does suggest these variants are not associated with the tumorigenesis in these individuals.

### Correlation of predicted HR status with histopathological markers

Breast tumor histological grade, ER status and TNBC status (negative for ER, PR and HER2) were previously shown as predictors of variant pathogenicity for classification of *BRCA1* or *BRCA2* germline variants in analysis of 4,477 *BRCA1* pathogenic variant carriers, 2,565 *BRCA2* pathogenic variant carriers, and 47,565 breast cancer cases without a known *BRCA1* or *BRCA2* variant.^46^ While number of observations in the dataset analyzed for our study is considerably smaller than the previous study,^46^ the overall trends in marker distribution were as expected. For example, based on the histopathology data available in our cohort (**Supplementary Table 4)**, ER-negative status was enriched in *BRCA1* positive individuals (22/25, 88%) compared to *BRCA2* positive individuals (2/21,10%) and individuals in the *BRCA1/2* negative group (40/149, 27%), which is comparable to trends reported previously for these groups (*BRCA1*, 76%; *BRCA2*, 21%; non-carrier, 23%).^46^ Similarly, the proportion of grade 3 tumors was highest for *BRCA1* positive individuals (19/25, 76%), intermediate for *BRCA2* positive individuals (12/21, 57%), and lowest for those in the *BRCA1/2* negative group (39/149, 26%), comparable to trends reported previously (*BRCA1*, 77%; *BRCA2*, 52%; non-carriers, 33%).^46^

The HR status and histopathological markers for each case, as well as age group and sex were visualized (**Supplementary Figure 5A**). Using the combined dataset from this study, there was a significant correlation between HR status by all tools and each of the histopathological markers, except for HER2 (**Supplementary Figure 5B**). For both CHORD and HRDetect, of all pathology markers, correlation was highest for TNBC status (r = 0.51), and lowest for HER2 status (r = 0.16-0.17). Further, there was a significant difference between TNBC and non-TNBC tumors in the distribution of the CHORD-predicted HRD probability (Wilcoxon p < 2.2e-16, **Supplementary Figure 5C**), and also *BRCA1* subtype predicted probability (Wilcoxon p = 6.5e-08, **Supplementary Figure 5D**).

## Discussion

Despite the strong known association of tumor HRD with *BRCA1/2*-associated hereditary cancer,^5^ and increasing use of tumor HRD as a biomarker for cancer treatment at the time of cancer diagnosis, this evidence type is not routinely used in germline variant classification of *BRCA1* and *BRCA2* genes. Our study leveraged data from four different cohorts (Familial breast, TCGA-BRCA, MAGIC, and Q-IMPROvE) of breast tumors from 350 individuals. Using HR status predicted with three different algorithms (CHORD, HRDetect, and HRDSum), we estimated the strength of evidence of breast tumor HR profile for predicting pathogenicity of *BRCA1* and *BRCA2* germline variants. We found HR status provides statistical justification for the potential utility of tumor HR profiling as an additional data source for *BRCA1* and *BRCA2* variant classification within existing specifications.^43^ Importantly, since different HR-calling algorithms are used in clinical practice, we investigated and demonstrated differences in the predictive capacity according to the HR testing method. We also assessed whether the prediction methods were robust for application to FFPE-derived samples.

The tumor material source for the MAGIC cohort was FFPE-derived samples, for which HR status prediction was unchanged after applying a single base substitution signature noise correction. This suggests that both HRDetect and CHORD prediction methods are robust for application to FFPE-derived samples in our cohort. However, since large-scale genome alterations such as structural and copy-number variations are a feature of HR prediction, we cannot rule out that the lower number of somatic SVs detected in the FFPE-derived tumor material from MAGIC may adversely impact HR prediction for some sample sets.

A particularly important outcome of this study is demonstrating the practical value of HR status for the assessment of *BRCA1/2* variant pathogenicity within existing guidelines. In the current *BRCA1/2* specifications, clinical data including breast tumor pathology status is currently captured within the ACMG/AMP PP4 and BP5 codes,^43^ with code weights dependent on the combined LR across tumor observations.^47^ To date, the breast tumor biomarkers used routinely in *BRCA1/2* variant classification include grade, ER status and TNBC status status.^46^ Calibrations from this previous study showed that most of the predictors were positively and negatively associated with *BRCA1* with supporting evidence strength level, with a few exceptions reaching moderate strength, while all of the predictions associated with *BRCA2* pathogenic germline variant status provided only pathogenic or benign supporting evidence for classification. Results from our study indicate that HR status is overall a stronger predictor of pathogenicity than other tumor pathology features currently used for both genes,^46^ for both pathogenic and benign directions. Evidence strengths applicable to HRD compared to HRP status were generally consistent for CHORD and HRDdetect, with evidence weights higher than when using HRDSum. This highlights the importance of validating HR prediction approaches within testing laboratories. The evidence towards and against pathogenicity associated with CHORD-predicted HRD subtypes reached pathogenic and benign strong strength for *BRCA1* and pathogenic and benign moderate strength for *BRCA2*, demonstrating that HR status is a more useful predictor than previously used tumor markers for variant classification. We also demonstrated that previously used breast tumor pathology markers (grade, ER and TNBC status) are correlated with HR status. Together these observations highlight the importance of selecting only one source of evidence derived from a given tumor for use in variant classification, to avoid double-counting. At this point in time, it would be logical to apply the evidence type providing the greatest weight (that is, HRD over e.g., grade).

Notably, separation by gene-specific HRD subtype predicted by CHORD can add another layer of precision, in that a VUS would not be assigned evidence towards pathogenicity if it was observed to have a tumor HRD profile with the opposite gene (e.g. a VUS in *BRCA2* within a *BRCA1*-like HRD tumor). This suggests that CHORD may provide more robust evidence towards pathogenicity for the classification of variants in *BRCA1* or *BRCA2* compared to other HR prediction algorithms. The gene-specific HRD subtype from CHORD invokes the exciting possibility that in the future, CHORD can be trained to predict non-*BRCA1/2* subtypes, such as *PALB2* or *RAD51C*. The ability of CHORD to predict which gene is associated with a HR deficient tumor will not only assist with classification of detected variants in these genes, but may also inform genetic analysis for patients with an undiagnosed germline cause of their cancer. For example, we hypothesize a patient with a HR deficient tumor predicted as *BRCA1*-like by CHORD, but with no pathogenic variants identified within the gene, may have variants within regulatory regions or promoter methylation that perturbs *BRCA1* and contributes to the HRD phenotype.^48^

This study also provides evidence weight based on HR predictions for an additional group of 21 VUS in *BRCA1/2*, and so may aid future classification of these variants. All except one variant were rare small indels or variants located in the 5’ or 3’ UTRs. These variant types are not well captured by existing classification guidelines, and thus this additional data could be beneficial to further inform classification.

It is worth emphasizing the importance of assessing the performance of the HR predictors dataset by dataset, to ensure that the calibration of predictors provides reliable results tailored to the specific characteristics of each dataset. This will include accounting for factors such as differences in HR measurement and/or HR status distribution between different tumor types, and between histological subtypes for a given tumor type. Larger studies will be important to validate our HR-associated LRs in other datasets, including reanalyzing the evidence weight associated with HR status by different age groups, clinical and pathology features. In particular, it will be necessary to re-investigate the value of HR status in other tumor types, such as ovarian cancer, for predicting germline variant pathogenicity, since predictive capacity is related to the prevalence of the tumor feature in individuals without a pathogenic germline variant. For example, while up to 80% of *BRCA1* or *BRCA2-*related ovarian cancers present with serous tumor histological subtype, this feature is not predictive of *BRCA1* or *BRCA2* variant pathogenicity since ∼70% of ovarian tumors without *BRCA1* or *BRCA2* pathogenic variants also present with this subtype.^20^

In summary, this study shows that relatively simple calibration approaches can be used to compare and select HR-calling algorithms for use in predicting pathogenicity of germline *BRCA1* and *BRCA2* variants, and that any HR predictor that is sufficiently predictive can be used to provide an alternative form of tumor data for application in germline *BRCA1* or *BRCA2* variant classification. Importantly, we have shown by comparison of different tumor HR prediction methods, our results have relevance for accurate detection of HR status, of clinical significance in guiding breast cancer treatment, since HR status is the trigger to ensure that patients might benefit from PARP inhibitor therapy.

## Data and code availability

The sequence data for the Familial breast cancers was previously deposited in the EGA under accession number <u>EGAD00001004494</u>. The TCGA data was from TCGA-BRCA and accessed from TCGA (https://portal.gdc.cancer.gov). The MAGIC dataset contains de-identified patient information using pseudo-identifiers to protect privacy. Please contact the corresponding authors for data access. The Q-IMPROvE data are available upon reasonable request to the authors Code and approaches used in this study: https://github.com/AdamaJava/adamajava and https://github.com/QingliGuo/FFPEsig.

## Data Availability

The sequence data for the Familial breast cancers was previously deposited in the EGA under accession number EGAD00001004494. The TCGA data was from TCGA-BRCA and accessed from TCGA (https://portal.gdc.cancer.gov). The MAGIC dataset contains de-identified patient information using pseudo-identifiers to protect privacy. Please contact the corresponding authors for data access. The Q-IMPROvE data are available upon reasonable request to the authors
Code and approaches used in this study: https://github.com/AdamaJava/adamajava and https://github.com/QingliGuo/FFPEsig.

## Acknowledgements

We are grateful to Prof Elgene Lim, St Vincent’s Sydney/Garvan; Dr Kate Cuff and Gillian Jagger, Princess Alexandra Hospital (PAH); Dr Kathryn Middleton, Mater Hospital South Brisbane; Dr Po-Ling Inglis and Dr Karin Steinke Royal Brisbane and Women’s Hospital (RBWH). We wish to thank Eveline Niedermayr, all the kConFab research nurses and staff, the heads and staff of the Family Cancer Clinics. kConFab is supported by a grant from the National Breast Cancer Foundation, and previously by the National Health and Medical Research Council (NHMRC), the Queensland Cancer Fund, the Cancer Councils of New South Wales, Victoria, Tasmania and South Australia, and the Cancer Foundation of Western Australia. We thank Dr Kate Cuff, Gillian Jagger, Dr Gorane Santamaria Hormaechea and the many staff across the RBWH, PAH and Mater hospitals and the Parkville Breast Unit and Familial Cancer Centre that helped facilitate this study. The results from the TCGA cohort are based upon data generated by the TCGA Research Network: https://www.cancer.gov/tcga. We also wish to acknowledge the Brisbane Breast Tissue Bank. Thank you to the patients and their families.

## Author contributions

**Cristina Fortuno:** Conceptualization, Investigation, Formal Analysis, Methodology, Writing – Original Draft Preparation. **Jia Zhang:** Investigation, Formal Analysis, Methodology, Visualisation, Data Curation, Writing – Original Draft Preparation. **Lambros T Koufariotis:** Formal Analysis, Resources, Writing - Review and Editing. **Georgina Hollway:** Methodology, Writing - Review and Editing. **Scott Wood:** Resources, Writing - Review and Editing. **John V Pearson:** Resources, Supervision, Writing - Review and Editing. **Peter Simpson:** Formal Analysis, Resources, Writing - Review and Editing. **Sunil R Lakhani:** Resources, Writing - Review and Editing. **Amy E McCart Reed:** Resources, Writing - Review and Editing. **Heather Thorne:** Data Curation, Resources, Writing - Review and Editing. **G Bruce Mann:** Resources, Writing - Review and Editing. **Anita R Skandarajah:** Resources, Writing - Review and Editing. **Lisa Devereux:** Resources, Writing - Review and Editing. **Qihong Zhao:** Resources, Writing - Review and Editing. **Dilanka L De Silva:** Resources, Writing - Review and Editing. **Geoffrey J Lindeman:** Resources, Supervision, Writing - Review and Editing. **Paul A James:** Conceptualization, Resources, Writing - Review and Editing. **Ian Campbell:** Resources, Supervision, Writing - Review and Editing, Funding Acquisition. **Amanda B Spurdle:** Conceptualization, Supervision, Project Administration, Methodology, Resources, Data Curation, Writing - Review and Editing, Funding Acquisition. **Nicola Waddell:** Conceptualization, Supervision, Project Administration, Methodology, Resources, Data Curation, Writing - Review and Editing, Funding Acquisition.

## Ethics statement

No participants were recruited for this study, the study included data from previously published or approved studies. The Q-IMPROVE was approved by a research ethics committee (HREC/2021/QRBW/73637). The MAGIC study received multisite institutional ethics approval from the Peter MacCallum Cancer Centre Human Research Ethics Committee (19/224, HREC/58844/PMCC-2019) and Governance approval obtained from each hospital site. Access to data and the analysis work in this study was approved by the QIMR Berghofer human ethics research committee within project number P2095, P2802 and P352

## References

1. Feng, C., Zhang, Y., Wu, F., Li, J., Liu, M., Lv, W., Li, C., Wang, W., Tan, Q., Xue, X., et al. (2023). Relationship between homologous recombination deficiency and clinical features of breast cancer based on genomic scar score. Breast 69, 392–400. 10.1016/j.breast.2023.04.002.

2. Turner, N.C. (2017). Signatures of DNA-Repair Deficiencies in Breast Cancer. N Engl J Med 377, 2490–2492. 10.1056/NEJMcibr1710161.

3. Wang, Z., Lu, Y., Han, M., Li, A., Ruan, M., Tong, Y., Yang, C., Zhang, X., Zhu, C., Wang, C., et al. (2024). Association between homologous recombination deficiency status and carboplatin treatment response in early triple-negative breast cancer. Breast cancer research and treatment 208, 429–440. 10.1007/s10549-024-07436-1.

4. Davies, H., Glodzik, D., Morganella, S., Yates, L.R., Staaf, J., Zou, X., Ramakrishna, M., Martin, S., Boyault, S., Sieuwerts, A.M., et al. (2017). HRDetect is a predictor of BRCA1 and BRCA2 deficiency based on mutational signatures. Nat Med 23, 517–525. 10.1038/nm.4292.

5. Creeden, J.F., Nanavaty, N.S., Einloth, K.R., Gillman, C.E., Stanbery, L., Hamouda, D.M., Dworkin, L., and Nemunaitis, J. (2021). Homologous recombination proficiency in ovarian and breast cancer patients. BMC cancer 21, 1154. 10.1186/s12885-021-08863-9.

6. Lee, J.E.A., Li, N., Rowley, S.M., Cheasley, D., Zethoven, M., McInerny, S., Gorringe, K.L., James, P.A., and Campbell, I.G. (2018). Molecular analysis of PALB2-associated breast cancers. J Pathol 245, 53–60. 10.1002/path.5055.

7. Nones, K., Johnson, J., Newell, F., Patch, A.M., Thorne, H., Kazakoff, S.H., de Luca, X.M., Parsons, M.T., Ferguson, K., Reid, L.E., et al. (2019). Whole-genome sequencing reveals clinically relevant insights into the aetiology of familial breast cancers. Annals of oncology: official journal of the European Society for Medical Oncology 30, 1071–1079. 10.1093/annonc/mdz132.

8. Prakash, R., Rawal, Y., Sullivan, M.R., Grundy, M.K., Bret, H., Mihalevic, M.J., Rein, H.L., Baird, J.M., Darrah, K., Zhang, F., et al. (2022). Homologous recombination-deficient mutation cluster in tumor suppressor RAD51C identified by comprehensive analysis of cancer variants. Proceedings of the National Academy of Sciences of the United States of America 119, e2202727119. 10.1073/pnas.2202727119.

9. Alexandrov, L.B., Nik-Zainal, S., Wedge, D.C., Aparicio, S.A., Behjati, S., Biankin, A.V., Bignell, G.R., Bolli, N., Borg, A., Børresen-Dale, A.L., et al. (2013). Signatures of mutational processes in human cancer. Nature 500, 415–421. 10.1038/nature12477.

10. Nik-Zainal, S., Alexandrov, L.B., Wedge, D.C., Van Loo, P., Greenman, C.D., Raine, K., Jones, D., Hinton, J., Marshall, J., Stebbings, L.A., et al. (2012). Mutational processes molding the genomes of 21 breast cancers. Cell 149, 979–993. 10.1016/j.cell.2012.04.024.

11. Nik-Zainal, S., Davies, H., Staaf, J., Ramakrishna, M., Glodzik, D., Zou, X., Martincorena, I., Alexandrov, L.B., Martin, S., Wedge, D.C., et al. (2016). Landscape of somatic mutations in 560 breast cancer whole-genome sequences. Nature 534, 47–54. 10.1038/nature17676.

12. Telli, M.L., Timms, K.M., Reid, J., Hennessy, B., Mills, G.B., Jensen, K.C., Szallasi, Z., Barry, W.T., Winer, E.P., Tung, N.M., et al. (2016). Homologous Recombination Deficiency (HRD) Score Predicts Response to Platinum-Containing Neoadjuvant Chemotherapy in Patients with Triple-Negative Breast Cancer. Clinical cancer research: an official journal of the American Association for Cancer Research 22, 3764–3773. 10.1158/1078-0432.Ccr-15-2477.

13. Nguyen, L. J. W.M.M., Van Hoeck, A., and Cuppen, E. (2020). Pan-cancer landscape of homologous recombination deficiency. Nature communications 11, 5584. 10.1038/s41467-020-19406-4.

14. Yao, H., Li, H., Wang, J., Wu, T., Ning, W., Diao, K., Wu, C., Wang, G., Tao, Z., Zhao, X., et al. (2023). Copy number alteration features in pan-cancer homologous recombination deficiency prediction and biology. Commun Biol 6, 527. 10.1038/s42003-023-04901-3.

15. Abbasi, A., Steele, C.D., Bergstrom, E.N., Khandekar, A., Farswan, A., McKay, R.R., Pillay, N., and Alexandrov, L.B. (2024). Detecting HRD in whole-genome and whole-exome sequenced breast and ovarian cancers. medRxiv. 10.1101/2024.07.14.24310383.

16. Yndestad, S., Engebrethsen, C., Herencia-Ropero, A., Nikolaienko, O., Vintermyr, O.K., Lillestøl, R.K., Minsaas, L., Leirvaag, B., Iversen, G.T., Gilje, B., et al. (2023). Homologous Recombination Deficiency Across Subtypes of Primary Breast Cancer. JCO Precis Oncol 7, e2300338. 10.1200/po.23.00338.

17. Purwar, R., Ranjan, R., Pal, M., Upadhyay, S.K., Kumar, T., and Pandey, M. (2023). Role of PARP inhibitors beyond BRCA mutation and platinum sensitivity in epithelial ovarian cancer: a meta-analysis of hazard ratios from randomized clinical trials. World J Surg Oncol 21, 157. 10.1186/s12957-023-03027-4.

18. Lotan, T.L., Kaur, H.B., Salles, D.C., Murali, S., Schaeffer, E.M., Lanchbury, J.S., Isaacs, W.B., Brown, R., Richardson, A.L., Cussenot, O., et al. (2021). Homologous recombination deficiency (HRD) score in germline BRCA2-versus ATM-altered prostate cancer. Mod Pathol 34, 1185–1193. 10.1038/s41379-020-00731-4.

19. Walsh, M.F., Ritter, D.I., Kesserwan, C., Sonkin, D., Chakravarty, D., Chao, E., Ghosh, R., Kemel, Y., Wu, G., Lee, K., et al. (2018). Integrating somatic variant data and biomarkers for germline variant classification in cancer predisposition genes. Human mutation 39, 1542–1552. 10.1002/humu.23640.

20. O’Mahony, D.G., Ramus, S.J., Southey, M.C., Meagher, N.S., Hadjisavvas, A., John, E.M., Hamann, U., Imyanitov, E.N., Andrulis, I.L., Sharma, P., et al. (2023). Ovarian cancer pathology characteristics as predictors of variant pathogenicity in BRCA1 and BRCA2. British journal of cancer 128, 2283–2294. 10.1038/s41416-023-02263-5.

21. Parsa, K., and Hasnain, S.E. (2015). Proteomics of multidrug resistant Mycobacterium tuberculosis clinical isolates: a peep show on mechanism of drug resistance & perhaps more. Indian J Med Res 141, 8–9. 10.4103/0971-5916.154485.

22. Fortuno, C., Mester, J., Pesaran, T., Weitzel, J.N., Dolinsky, J., Yussuf, A., McGoldrick, K., Garber, J.E., Savage, S.A., Khincha, P.P., et al. (2020). Suggested application of HER2+ breast tumor phenotype for germline TP53 variant classification within ACMG/AMP guidelines. Human mutation. 10.1002/humu.24060.

23. Thompson, B.A., Goldgar, D.E., Paterson, C., Clendenning, M., Walters, R., Arnold, S., Parsons, M.T., Michael, D.W., Gallinger, S., Haile, R.W., et al. (2013). A multifactorial likelihood model for MMR gene variant classification incorporating probabilities based on sequence bioinformatics and tumor characteristics: a report from the Colon Cancer Family Registry. Human mutation 34, 200–209. 10.1002/humu.22213.

24. Thorne, H., Mitchell, G., and Fox, S. (2011). kConFab: a familial breast cancer consortium facilitating research and translational oncology. J Natl Cancer Inst Monogr 2011, 79–81. 10.1093/jncimonographs/lgr042.

25. Thennavan, A., Beca, F., Xia, Y., Recio, S.G., Allison, K., Collins, L.C., Tse, G.M., Chen, Y.Y., Schnitt, S.J., Hoadley, K.A., et al. (2021). Molecular analysis of TCGA breast cancer histologic types. Cell Genom 1. 10.1016/j.xgen.2021.100067.

26. De Silva, D.L., Stafford, L., Skandarajah, A.R., Sinclair, M., Devereux, L., Hogg, K., Kentwell, M., Park, A., Lal, L., Zethoven, M., et al. (2023). Universal genetic testing for women with newly diagnosed breast cancer in the context of multidisciplinary team care. Med J Aust 218, 368–373. 10.5694/mja2.51906.

27. McCart Reed, A.E., Hollway, G., and Lakhani, S. (2023). The Queensland IMplementation of PRecision Oncology in brEast cancer (Q-IMPROvE) pilot study. Med J Aust 218, 374–375. 10.5694/mja2.51900.

28. Li, H., and Durbin, R. (2009). Fast and accurate short read alignment with Burrows-Wheeler transform. Bioinformatics 25, 1754–1760. 10.1093/bioinformatics/btp324.

29. Kassahn, K.S., Holmes, O., Nones, K., Patch, A.M., Miller, D.K., Christ, A.N., Harliwong, I., Bruxner, T.J., Xu, Q., Anderson, M., et al. (2013). Somatic point mutation calling in low cellularity tumors. PloS one 8, e74380. 10.1371/journal.pone.0074380.

30. Poplin, R., Ruano-Rubio, V., DePristo, M.A., Fennell, T.J., Carneiro, M.O., Van der Auwera, G.A., Kling, D.E., Gauthier, L.D., Levy-Moonshine, A., Roazen, D., et al. (2018). Scaling accurate genetic variant discovery to tens of thousands of samples. bioRxiv, 201178. 10.1101/201178.

31. Cingolani, P., Platts, A., Wang le, L., Coon, M., Nguyen, T., Wang, L., Land, S.J., Lu, X., and Ruden, D.M. (2012). A program for annotating and predicting the effects of single nucleotide polymorphisms, SnpEff: SNPs in the genome of Drosophila melanogaster strain w1118; iso-2; iso-3. Fly (Austin) *6*, 80-92. 10.4161/fly.19695.

32. Raine, K.M., Van Loo, P., Wedge, D.C., Jones, D., Menzies, A., Butler, A.P., Teague, J.W., Tarpey, P., Nik-Zainal, S., and Campbell, P.J. (2016). ascatNgs: Identifying Somatically Acquired Copy-Number Alterations from Whole-Genome Sequencing Data. Curr Protoc Bioinformatics 56, 15.19.11-15.19.17. 10.1002/cpbi.17.

33. Hayward, N.K., Wilmott, J.S., Waddell, N., Johansson, P.A., Field, M.A., Nones, K., Patch, A.M., Kakavand, H., Alexandrov, L.B., Burke, H., et al. (2017). Whole-genome landscapes of major melanoma subtypes. Nature 545, 175–180. 10.1038/nature22071.

34. Guo, Q., Lakatos, E., Bakir, I.A., Curtius, K., Graham, T.A., and Mustonen, V. (2022). The mutational signatures of formalin fixation on the human genome. Nature communications 13, 4487. 10.1038/s41467-022-32041-5.

35. Abkevich, V., Timms, K.M., Hennessy, B.T., Potter, J., Carey, M.S., Meyer, L.A., Smith-McCune, K., Broaddus, R., Lu, K.H., Chen, J., et al. (2012). Patterns of genomic loss of heterozygosity predict homologous recombination repair defects in epithelial ovarian cancer. British journal of cancer 107, 1776–1782. 10.1038/bjc.2012.451.

36. Popova, T., Manié, E., Rieunier, G., Caux-Moncoutier, V., Tirapo, C., Dubois, T., Delattre, O., Sigal-Zafrani, B., Bollet, M., Longy, M., et al. (2012). Ploidy and large-scale genomic instability consistently identify basal-like breast carcinomas with BRCA1/2 inactivation. Cancer research 72, 5454–5462. 10.1158/0008-5472.Can-12-1470.

37. Birkbak, N.J., Wang, Z.C., Kim, J.Y., Eklund, A.C., Li, Q., Tian, R., Bowman-Colin, C., Li, Y., Greene-Colozzi, A., Iglehart, J.D., et al. (2012). Telomeric allelic imbalance indicates defective DNA repair and sensitivity to DNA-damaging agents. Cancer Discov 2, 366–375. 10.1158/2159-8290.Cd-11-0206.

38. Sztupinszki, Z., Diossy, M., Krzystanek, M., Reiniger, L., Csabai, I., Favero, F., Birkbak, N.J., Eklund, A.C., Syed, A., and Szallasi, Z. (2018). Migrating the SNP array-based homologous recombination deficiency measures to next generation sequencing data of breast cancer. NPJ Breast Cancer 4, 16. 10.1038/s41523-018-0066-6.

39. Ray-Coquard, I., Pautier, P., Pignata, S., Pérol, D., González-Martín, A., Berger, R., Fujiwara, K., Vergote, I., Colombo, N., Mäenpää, J., et al. (2019). Olaparib plus Bevacizumab as First-Line Maintenance in Ovarian Cancer. N Engl J Med 381, 2416–2428. 10.1056/NEJMoa1911361.

40. Liu, X., Li, C., Mou, C., Dong, Y., and Tu, Y. (2020). dbNSFP v4: a comprehensive database of transcript-specific functional predictions and annotations for human nonsynonymous and splice-site SNVs. Genome Med 12, 103. 10.1186/s13073-020-00803-9.

41. Landrum, M.J., Lee, J.M., Benson, M., Brown, G.R., Chao, C., Chitipiralla, S., Gu, B., Hart, J., Hoffman, D., Jang, W., et al. (2018). ClinVar: improving access to variant interpretations and supporting evidence. Nucleic Acids Res 46, D1062–D1067. 10.1093/nar/gkx1153.

42. Chen, S., Francioli, L.C., Goodrich, J.K., Collins, R.L., Kanai, M., Wang, Q., Alföldi, J., Watts, N.A., Vittal, C., Gauthier, L.D., et al. (2024). A genomic mutational constraint map using variation in 76,156 human genomes. Nature 625, 92–100. 10.1038/s41586-023-06045-0.

43. Parsons, M.T., de la Hoya, M., Richardson, M.E., Tudini, E., Anderson, M., Berkofsky-Fessler, W., Caputo, S.M., Chan, R.C., Cline, M.S., Feng, B.J., et al. (2024). Evidence-based recommendations for gene-specific ACMG/AMP variant classification from the ClinGen ENIGMA BRCA1 and BRCA2 Variant Curation Expert Panel. Am J Hum Genet 111, 2044–2058. 10.1016/j.ajhg.2024.07.013.

44. Tavtigian, S.V., Greenblatt, M.S., Harrison, S.M., Nussbaum, R.L., Prabhu, S.A., Boucher, K.M., and Biesecker, L.G. (2018). Modeling the ACMG/AMP variant classification guidelines as a Bayesian classification framework. Genetics in medicine: official journal of the American College of Medical Genetics. 10.1038/gim.2017.210.

45. Tavtigian, S.V., Harrison, S.M., Boucher, K.M., and Biesecker, L.G. (2020). Fitting a naturally scaled point system to the ACMG/AMP variant classification guidelines. Human mutation. 10.1002/humu.24088.

46. Spurdle, A.B., Couch, F.J., Parsons, M.T., McGuffog, L., Barrowdale, D., Bolla, M.K., Wang, Q., Healey, S., Schmutzler, R., Wappenschmidt, B., et al. (2014). Refined histopathological predictors of BRCA1 and BRCA2 mutation status: a large-scale analysis of breast cancer characteristics from the BCAC, CIMBA, and ENIGMA consortia. Breast cancer research: BCR 16, 3419. 10.1186/s13058-014-0474-y.

47. Parsons, M.T., Tudini, E., Li, H., Hahnen, E., Wappenschmidt, B., Feliubadalo, L., Aalfs, C.M., Agata, S., Aittomaki, K., Alducci, E., et al. (2019). Large scale multifactorial likelihood quantitative analysis of BRCA1 and BRCA2 variants: An ENIGMA resource to support clinical variant classification. Human mutation. 10.1002/humu.23818.

48. Schwartz, M., Ibadioune, S., Delhomelle, H., Barraud, S., Caputo, S.M., Trabelsi-Grati, O., Villy, M.C., Laugé, A., Tang, R., Rouleau, E., et al. (2025). High prevalence of constitutional BRCA1 epimutation in patients with early-onset triple-negative breast cancer. Clinical epigenetics 17, 91. 10.1186/s13148-025-01885-1.

