## Supplementary Figures for "Integrating breast tumor homologous recombination deficiency status to aid germline *BRCA1* and *BRCA2* variant classification"

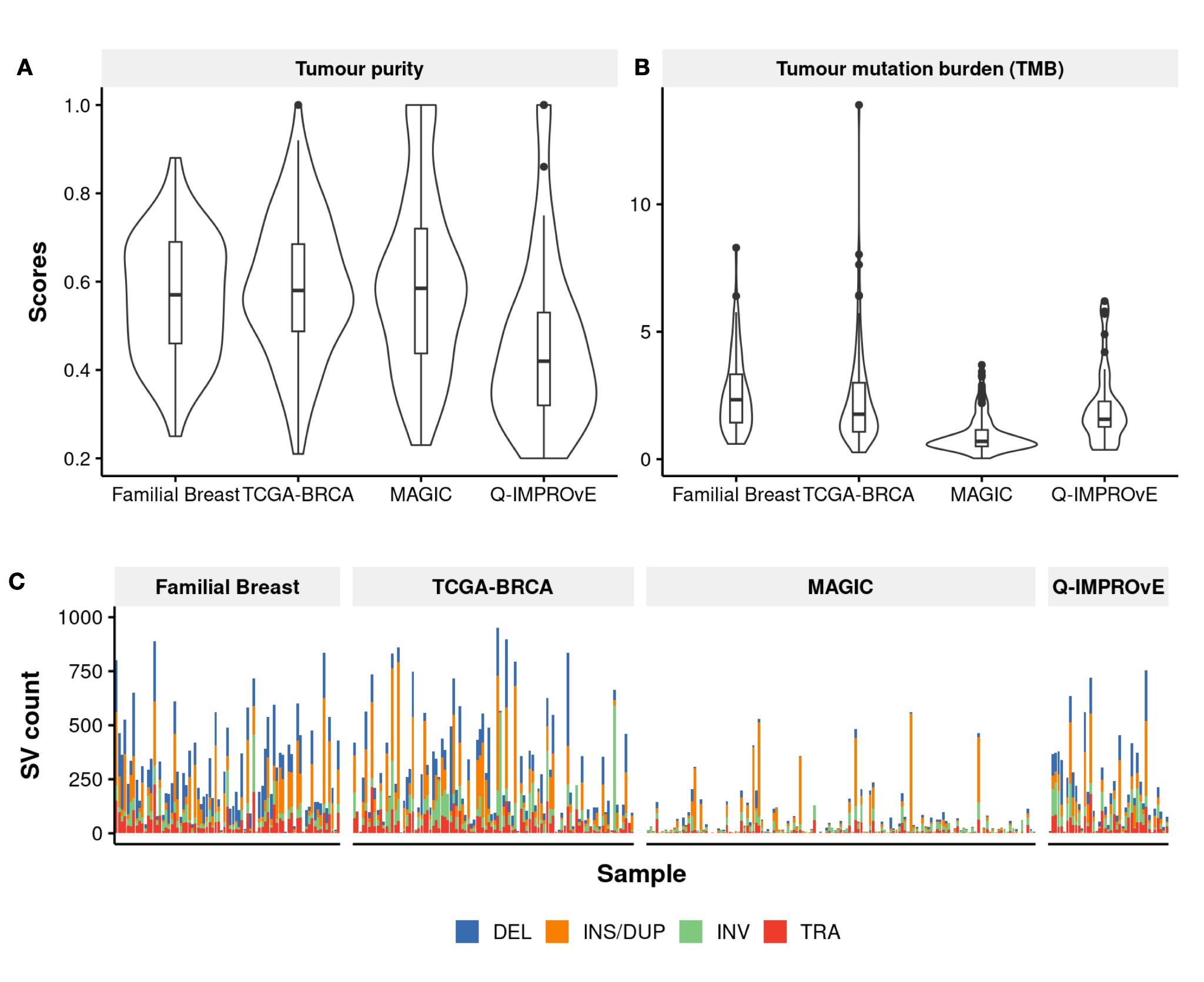

**Supplementary Figure 1. Tumor purity and tumor mutation burden (TMB) at nonsynonymous sites and SV count in each cohort.** A) Violin and boxplots display the distribution of tumor purity scores estimated using ascatNGS. B) Violin and box plots for the TMB at nonsynonymous sites. C) The number of somatic SVs in samples across four cohorts, with each bar colored by SV types (DEL: blue, INS/DUP: orange, INV: green, TRA: red). Samples in each panel are stratified by cohort and include Familial Breast (n=77), TCGA-BRCA (n=96), MAGIC (n=136) and Q-IMPROvE (n=41)

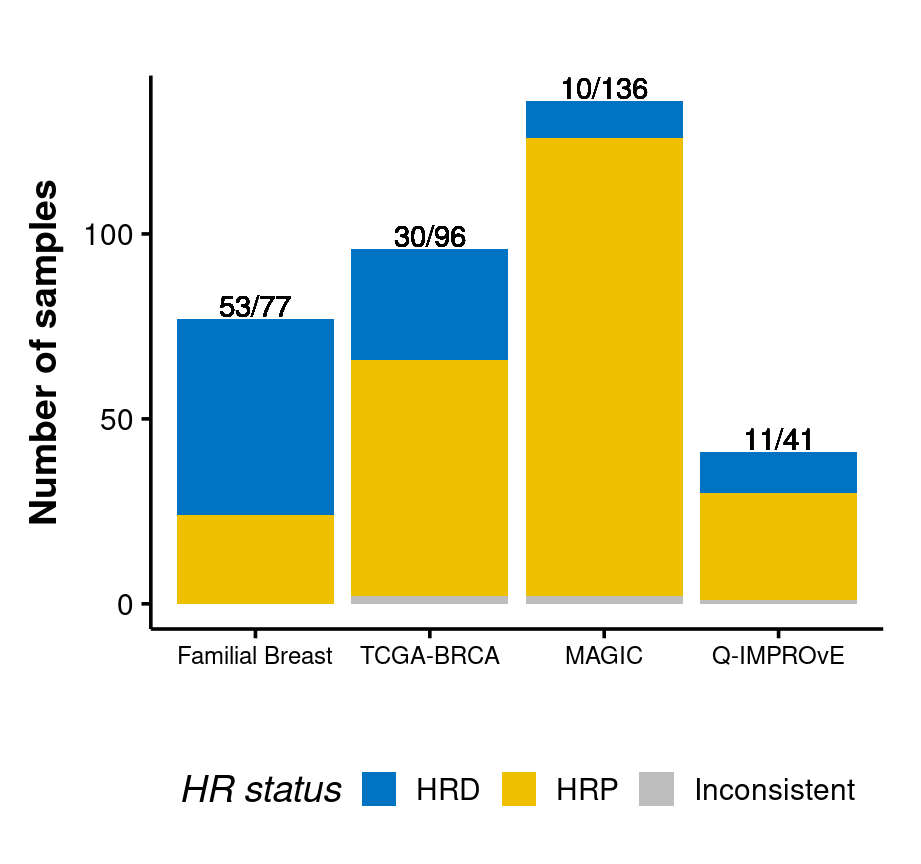

**Supplementary Figure 2. Barplot displays the number of samples identified as HRD or HRP by CHORD and HRDetect for four cohorts.** The consistent CHORD and HRDetect predictions were colored by HR status (blue: HRD, yellow: HRP), and inconsistent predictions were colored in grey. The number of HRD samples of each cohort was labelled at the top of each bar

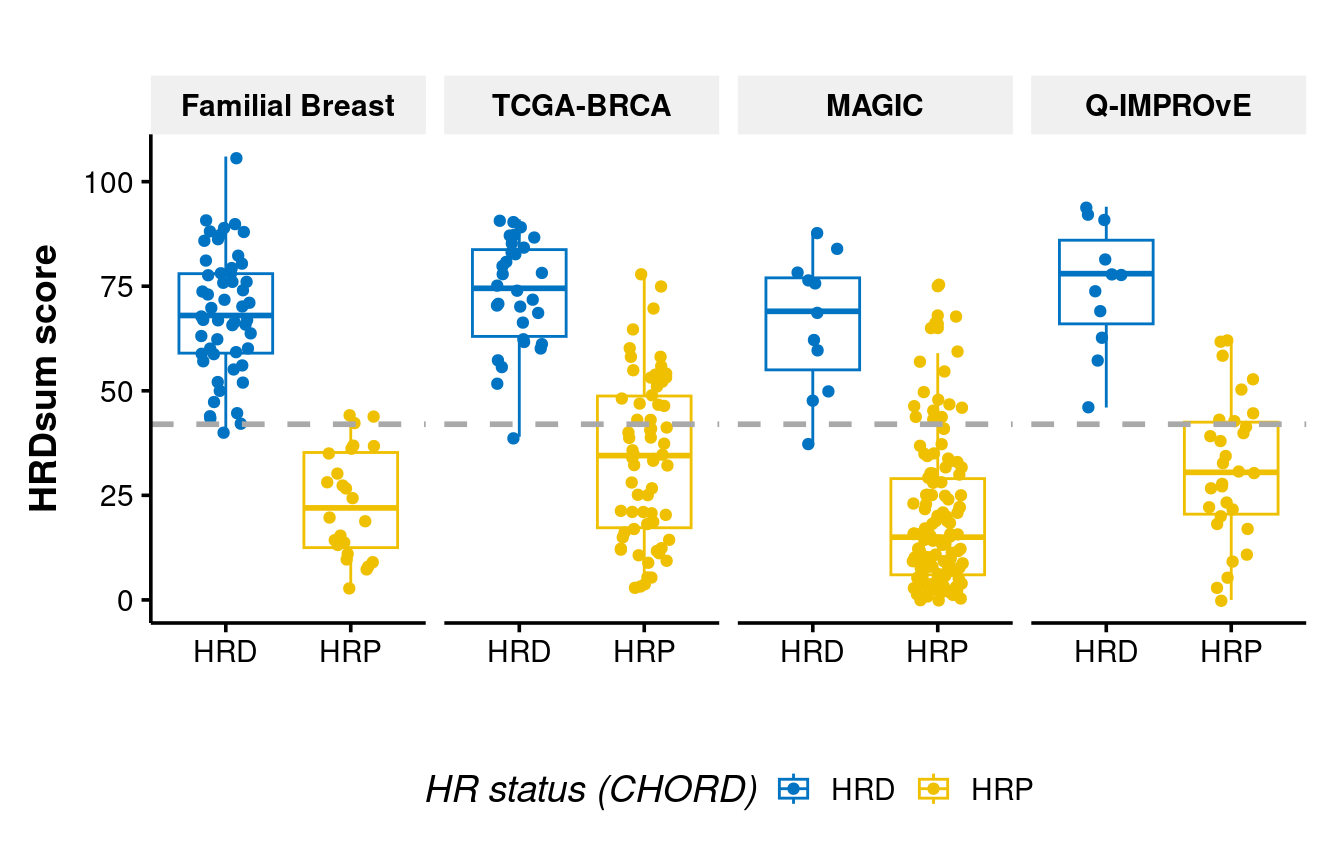

**Supplementary Figure 3**. **Distribution of HRDSum scores across the four cohorts.** HRDsum scores of all samples were stratified as HRD and HRP based on the CHORD prediction. Each boxplot displays the distribution of HRDsum scores within each group. The box represents the interquartile range (IQR; 25th–75th percentile), with the horizontal line inside indicating the median. Whiskers extend to the most extreme values within 1.5× IQR from the quartiles. The grey dashed line denotes the HRDsum threshold of 42 used to classify samples

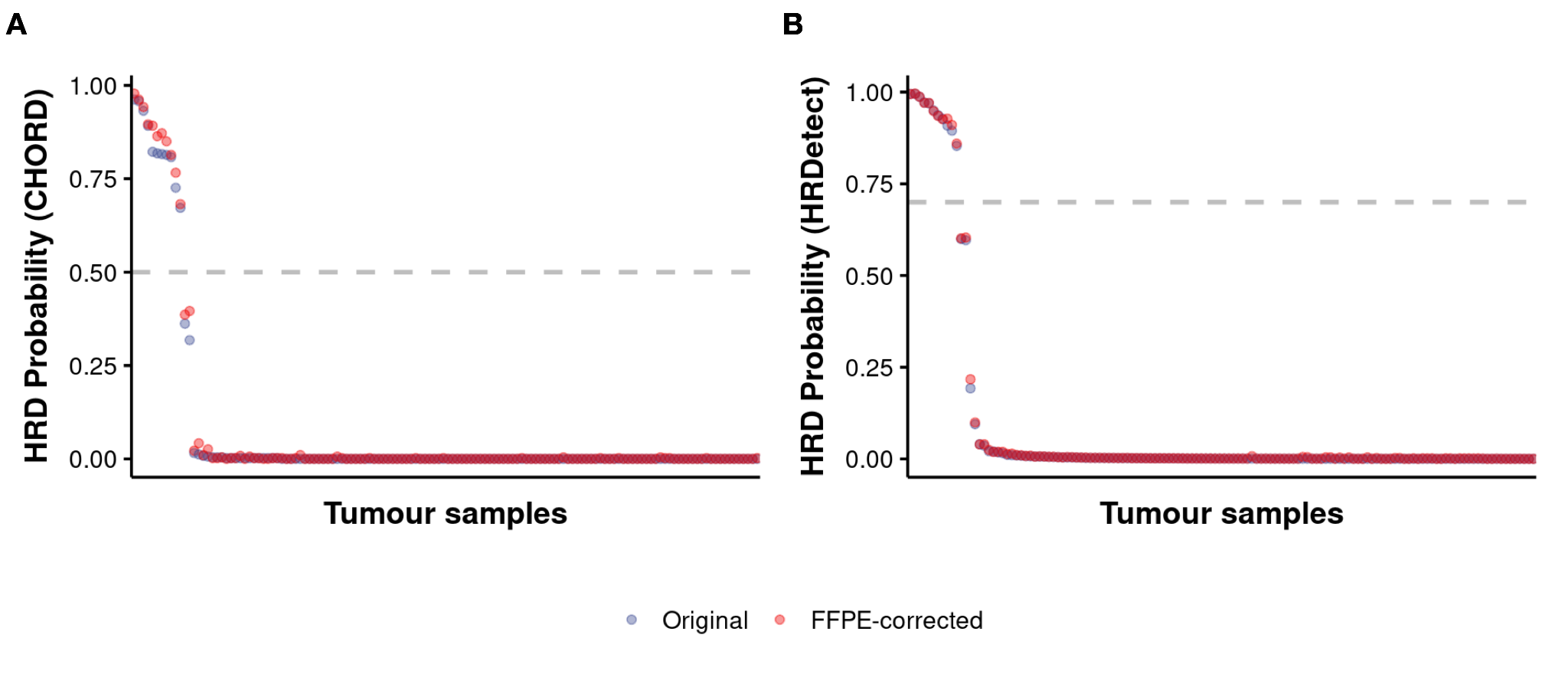

**Supplementary Figure 4. Comparison of HRD prediction results before and after FFPE signature correction.** Points represent the HRD prediction scores for each sample using CHORD (A) or HRDetect (B). The original HRD prediction scores were colored in blue and scores after correction were in red. The thresholds for CHORD and HRDetect are shown in the plot with grey dashed lines

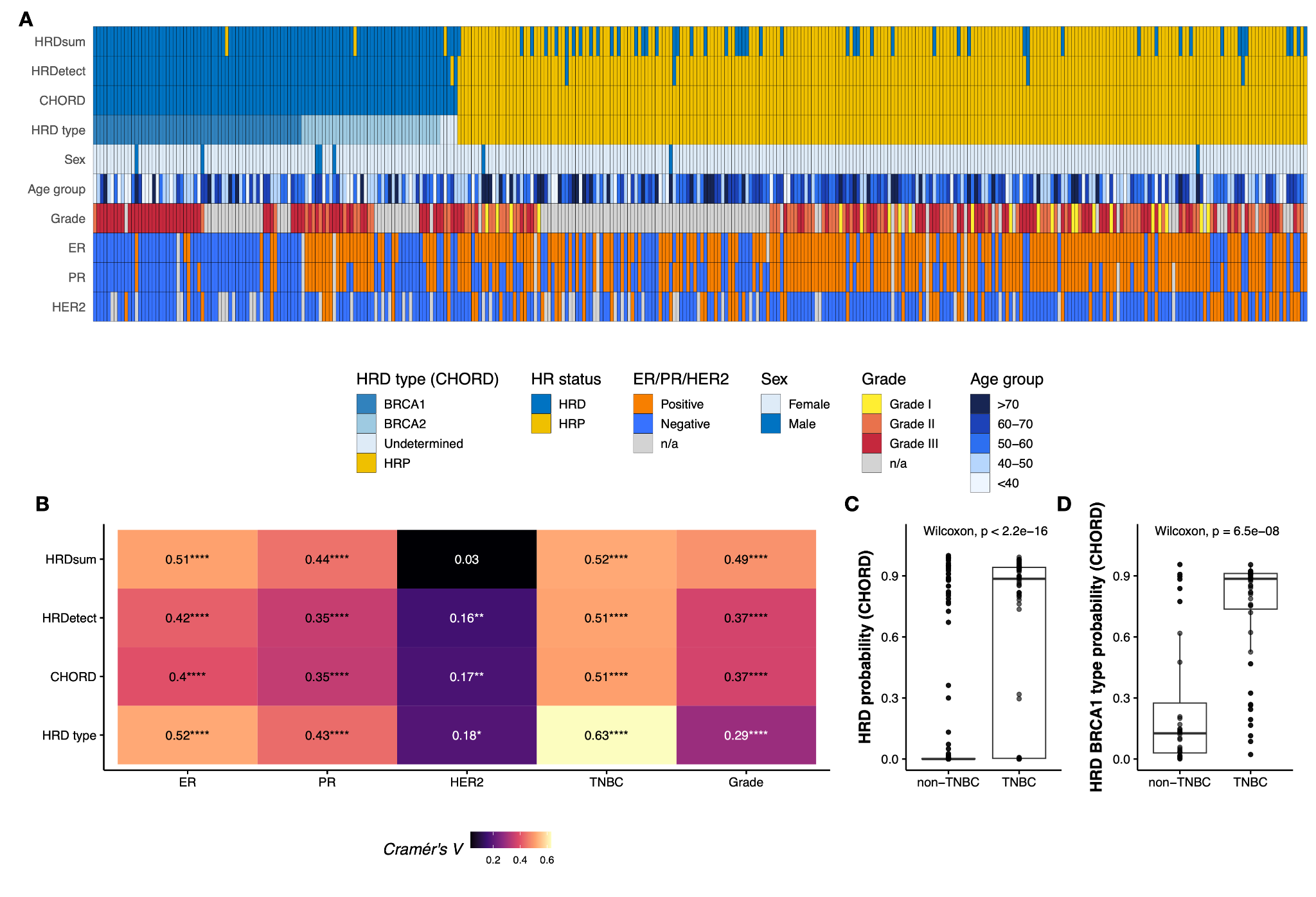

**Supplementary Figure 5. The correlation of sample pathological information with the HR status prediction.** A) HR status prediction from HRDsum, HRDetect, and CHORD, and clinical information for each sample, including sex, age group, tumor histological grade, hormone biomarker status (ER, PR, HER2). B) Associations between HR status predictions (from HRDsum, HRDetect, CHORD) and clinicopathological variables (ER, PR, HER2 status, TNBC status, and histological grade). CHORD-predicted HRD subtypes are also shown. Statistical significance was assessed using the Chi-square test and indicated as follows: P < 0.05 (*), P < 0.01 (**), P < 0.001 (***), P < 0.0001 (****). P < 0.0025 (*** and ****) is considered significant after Bonferroni correction for multiple testing. Cramér’s V values show the strength of association, with brighter colors denoting stronger correlations. C) HRD probability scores from CHORD for samples grouped by TNBC and non-TNBC status. P value using the Wilcoxon rank-sum test. D) HRD BRCA1 type from CHORD for samples grouped by TNBC and non-TNBC status. P-value using the Wilcoxon rank-sum test
